# Subjective sleep quality and objective sleep physiology in migraineurs: a meta-analysis

**DOI:** 10.1101/2021.03.03.21252791

**Authors:** E.C. Stanyer, H. Creeney, A.D. Nesbitt, P. R. Holland, J. Hoffmann

## Abstract

**Objectives:** Sleep disturbance is often associated with migraine. However, there is a paucity of research investigating objective and subjective measures of sleep in migraineurs. This meta-analysis aims to determine whether there are differences in subjective sleep quality measured using the Pittsburgh Sleep Quality Index (PSQI) and objective sleep physiology measured using polysomnography between adult and pediatric migraineurs, and healthy controls.

**Methods:** A systematic search of five databases was conducted to find case-controlled studies which measured polysomnography and/or PSQI in migraineurs. Effect sizes (Hedges’ *g*) were entered into a random effects model meta-analysis.

**Results:** 32 separate studies were eligible. Overall, adult migraineurs had higher PSQI scores than healthy controls (*g* = 0.75, *p* < .001). This effect was larger in chronic than episodic migraineurs (*g* = 1.03, *p* < .001, *g* = 0.63, *p* < .001 respectively). For polysomnographic studies, adult and pediatric migraineurs displayed a lower percentage of REM sleep (*g* = −0.22, *p* = 0.017, *g* = −0.71, *p* = 0.025 respectively) than healthy controls. Pediatric migraineurs also displayed less total sleep time (*g* = −1.37, *p* = 0.039), more wake (*g* = 0.52, *p* < .001) and shorter sleep onset latency (*g* = −0.37, *p* < .001) than healthy controls.

**Conclusion:** Migraineurs have significantly poorer subjective sleep quality, and altered sleep compared to healthy individuals – a finding which is particularly evident in children. This has implications for developing appropriate treatments. Further longitudinal empirical studies are required to enhance our understanding of this relationship.

## 1. Introduction

It has long been recognized that there is a relationship between sleep and migraine. This relationship is complex, as alterations in sleep can be a trigger, treatment, or symptom of migraine^1^. There are an estimated one billion migraineurs globally, and migraine is one of the leading causes of disability worldwide^2^, with a considerable personal and socioeconomic burden^3^. To reduce this burden and meet the growing clinical need^4^, a clearer understanding of the profile of sleep in migraineurs and its relation to migraine-related disability is important, to enable clinicians to support migraineurs and deliver targeted, effective sleep interventions^5^.

Despite its association there remains a paucity of research into sleep in migraineurs, and there is no consensus on whether migraineurs exhibit objective changes in sleep physiology. This is partly due to the small sample sizes of polysomnographic (PSG) studies used to measure sleep. The current meta-analysis aims to overcome this by aggregating data from multiple studies investigating differences in subjective sleep quality as measured by the Pittsburgh Sleep Quality Index (PSQI)^6^, and objective sleep physiology measured using PSG between migraineurs and healthy controls. Furthermore, the relationship between measures of subjective sleep quality and migraine disability was investigated by combining correlational data between PSQI and Migraine Disability Assessment Test (MIDAS) scores^7^.

## 2. Methods

The format of this review followed the Preferred Reporting Items for Systematic Reviews and Meta-analysis (PRISMA) guidelines^8^ and the PRISMA 2009 checklist (Table e-1). The protocol for this review was pre-registered with PROSPERO (registration number: CRD42020209325).

### 2.1. Search Strategy

Two authors (ECS and HC) conducted an independent, electronic search of relevant databases [Embase (1996 - 2020), MEDLINE® (1996 - 2020), Global Health (1973 – 2020), and APA PsycINFO (1806 - 2020), APA PsycArticles Full Text (2020)] from their inception to the current date. Key search terms included combinations of *migraine, sleep*, PSQI, Pittsburgh Sleep Quality Index, polysomnograph*, PSG, EEG, electroencephalograph*, MIDAS, HIT-6, MSQ* with Boolean operators. The full search strategy for OVID is displayed in Table e-2. The search was limited to studies published in English and duplicates were removed. Titles and abstracts were independently screened for eligibility by two authors (ECS and HC). Studies that were eligible or if eligibility was unclear were submitted to the full text review stage. Relevant studies were also retrieved from the reference lists of eligible studies. All full texts were screened for eligibility by one reviewer (ECS) and 10% of the full texts were selected randomly using a random number generator which the second reviewer (HC) then screened for eligibility. Any discrepancies were resolved with the aid of another reviewer. All studies were examined to ensure they were independent of one another. If full texts were not available, the original authors were contacted, and the full texts were requested. The last search date was the 17th of December 2020.

### 2.2. Inclusion Criteria

Studies were included which examined sleep quality as assessed by the PSQI and/or physiological sleep variables using PSG in adult and pediatric migraineurs and healthy control participants. Studies which computed correlations between MIDAS and PSQI scores in migraineurs were also included. Review articles or single-case studies were not included. Studies which did not have a suitable healthy control group were excluded.

### 2.3. Population

There were no restrictions on the age of the participants in the studies. However, for analysis purposes, adults ≥18 years of age and children <18 years of age were included in separate analyses given that sleep physiology demonstrates age-dependent quantitative differences^9^. Pregnant participants, and participants with other headache disorders namely: cluster headache, tension type headache, and medication over-use headache were not included in the analysis. However, a study was included if it reported data which could be extracted that was specific to migraineurs and no other headache disorders. Due to the limited number of studies in this area, we deliberately kept the classification of migraine broad, thus migraineurs with any diagnosis were included in this analysis: episodic, chronic, migraine without aura (MO), migraine with aura (MA). Although this is not a standard definition in the International Classification of Headache Disorders-3, studies which also categorized their patients as sleep-related migraine (SRM), or non-sleep-related migraine (NSRM) were included. Studies which categorized their patients by the number of migraine or headache days per month, and not by episodic or chronic were also included. In such cases, we categorized patients experiencing headache/migraine on ≥15 days per month as chronic and pooled them with the other studies in chronic migraineurs. As sleep quality may differ between patients with different frequencies of migraine, episodic and chronic migraine were analyzed as subgroups in the meta-analysis of PSQI scores.

### 2.4. Outcome Measures

To indicate whether migraineurs experience altered sleep, the primary outcomes calculated were weighted effect sizes (Hedges’ *g*) for the difference between migraineurs and controls in global PSQI score and PSG-derived sleep measures. The PSG-derived sleep measures were total sleep time (TST) in minutes, sleep efficiency percent (SE), percent wake, percent of TST spent in REM sleep, non-REM (NREM) sleep stage 1 (N1), stage 2 (N2), and stage 3 (N3), sleep onset latency (SOL) in minutes. Secondly, to ascertain whether subjective sleep quality is related to migraine disability the Fisher’s *z* transformed correlation coefficient between MIDAS and PSQI scores in the migraine population was calculated.

### 2.5. Data Extraction

Data extraction was performed using an a priori elaborated table in Microsoft Excel. Extraction was completed by one author (ECS) and included: authors, year of publication, journal, publication type, database extracted from, participant demographics (mean age, sex ratio), migraine characterization, mean global PSQI score, PSG-derived sleep variable means, correlation coefficient between MIDAS and PSQI scores, standard deviations for each data point, and sample size. Where the measures were not reported directly in the publication or the data were not in the correct format for analysis (e.g., medians instead of means), the authors were contacted to request this data. Studies which did not report data from which the effect sizes could be calculated after contacting the authors, or if the authors did not respond were excluded. Further data were extracted that might be potential moderators where appropriate. For example: the design of the study (e.g., matched, or non-matched controls), whether the study excluded those on medication which may affect the sleep cycle or those with co-morbid sleep disorders, and presence of a PSG adaptation night or not. If studies used older sleep scoring criteria to determine NREM sleep stages separately (stage 3 and stage 4 sleep), an average of the means for these two stages was computed to be comparable to updated AASM nomenclature: which defines these singularly as N3. If a study reported PSG sleep stage variables in minutes rather than percentages these were calculated manually based on the TST means available.

### 2.6. Statistical Analysis

Statistical analysis was performed using RStudio and the *metafor* package. The weighted effect sizes (Hedges’ *g*) for each study were calculated using the means, sample size (*n*) and standard deviations for each of the measures. Effect sizes were interpreted as small (0.2), medium (0.5) or large (0.8) in line with guidelines and were visualized using forest plots. The effect sizes were calculated such that a negative Hedges’ *g* value indicated that the controls had a higher score on that measure. For the PSQI analysis, studies which investigated chronic migraineurs were initially pooled with episodic migraineurs for a global analysis of effect size, however they were then analyzed in two separate sub-groups. For the sleep physiology analysis, PSG-derived variables from pediatric and adult migraineurs were analyzed as two separate groups. Effect sizes were computed for each PSG measure separately.

#### 2.6.1. Publication bias

Publication bias refers to the over-inflation of effect sizes due to the tendency for non-significant findings to remain unpublished. To assess this, Egger’s regression test for funnel plot asymmetry was performed. If the result is significant at *p* < 0.05, this provides evidence of publication bias. However, as Egger’s test is prone to producing false positives particularly with small numbers of studies, publication bias was also assessed by visual inspection of funnel plots. These plot a measure of precision (standard error) against the observed effect size (Hedges’ *g*). If the funnel plots are substantially asymmetric, then publication bias can be assumed. Duval and Tweedie’s trim-and-fill method was used to assess whether there were any unpublished studies missing from the analysis and estimate what the adjusted effect size would be if these studies were present.

#### 2.6.2. Between-studies heterogeneity

As a meta-analysis typically includes studies of varying designs and populations, it is important to quantify the proportion of between-studies heterogeneity to ensure accurate estimation of the summary effect sizes. Cochran’s *Q* statistic is commonly used to assess this. If *Q* is significant at *p* < 0.05 this indicates variability in the effect sizes reported between studies. However, as this test has poor power to detect heterogeneity when the meta-analysis includes a small number of studies the *I*^2^ statistic was also calculated. An *I*^2^ value of 0% represents no heterogeneity, 25% low, 50% moderate and 75% high. As heterogeneity was to be expected given the variation in participant characteristics and experimental designs, a random-effects model meta-analysis was conducted for all measures.

To explore potential sources of heterogeneity in both the PSQI and PSG analyses, where a significant effect size and moderate heterogeneity as indexed by an *I*^2^ of >50% were found, additional analyses were conducted by including characteristics of the study as moderator variables. The variables which were included as moderators were not predefined and were based on a previous meta-analysis^10^: exclusion of participants with sleep disorders (yes/no), exclusion of participants taking drugs which affect the sleep cycle or a washout period before the study (yes/no), whether the control population was matched for sex and age (yes/no). Where a study did not state in the paper whether this was conducted or not, it was coded as ‘no’ for the purposes of analysis.

### 2.7. Data Availability Statement

The source data are available from the individual studies included. The extracted raw data are available at Open Science Framework: https://osf.io/3t4u5/.

## 3. Results

### 3.1. Description of Studies

Results of the study selection process are displayed in **Figure 1**. From a total of 4089 studies after duplicates were removed, 183 were identified as potentially eligible after screening the titles and abstracts. The full texts were then screened to confirm this, after which 32 independent studies were included in the meta-analysis. Table 1 presents an overview of the studies included in the review. Twenty investigated sleep quality in adults with and without migraine using the PSQI. Of these, 14 of them had a population consisting of episodic migraineurs and 6 of them chronic migraineurs. Four studies (two already included in the PSQI analysis) reported the correlation between MIDAS and PSQI scores in migraineurs^11–14^. There were no participants <18 years of age in the PSQI analysis or the MIDAS and PSQI correlational analysis due to a lack of available data. Eleven studies measured PSG in adults^15–20^ and children^21–25^, with and without migraine. One study^16^ compared migraineurs PSG in the pre-, mid- and post-ictal phase, and thus it was not appropriate to extract a single value for this data. However, they provided a pooled value for the PSQI analysis, and thus it was excluded from the PSG analysis only.

**Figure 1:**
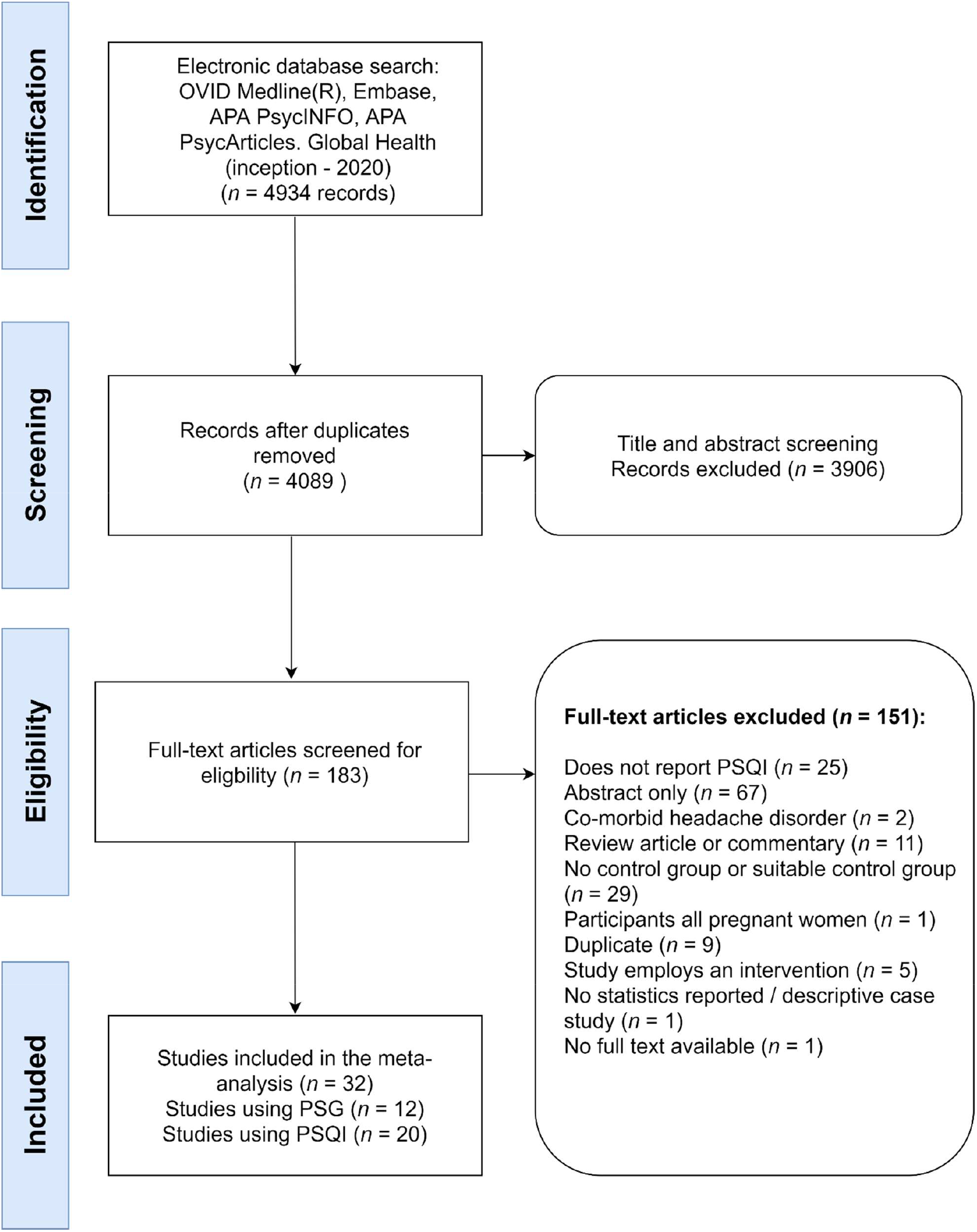
PRISMA flow diagram depicting the stages of study selection. Abbreviations: PSQI, Pittsburgh Sleep Quality Index; PSG, polysomnography; MIDAS, Migraine Disability Assessment.

**Table 1:**
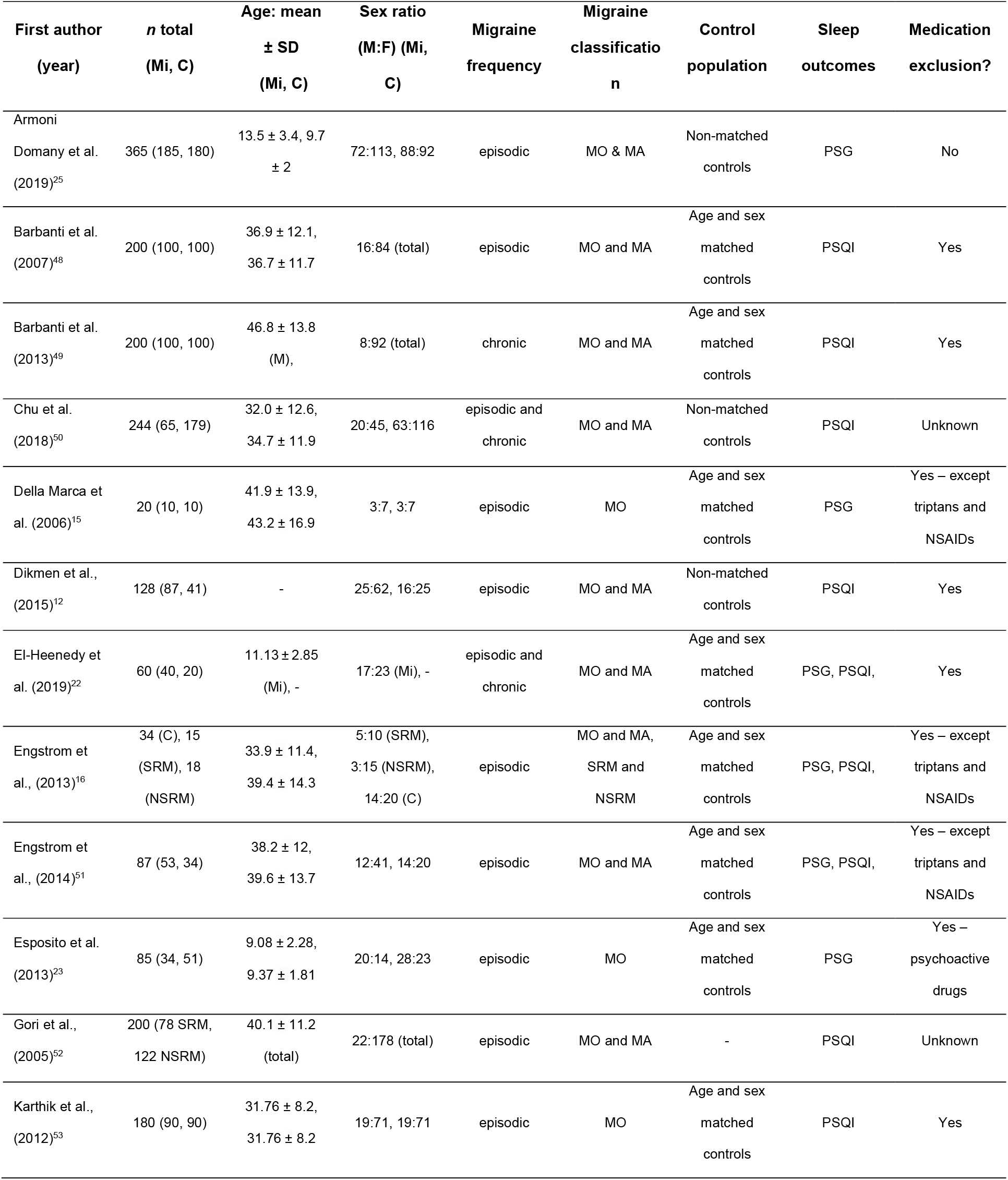
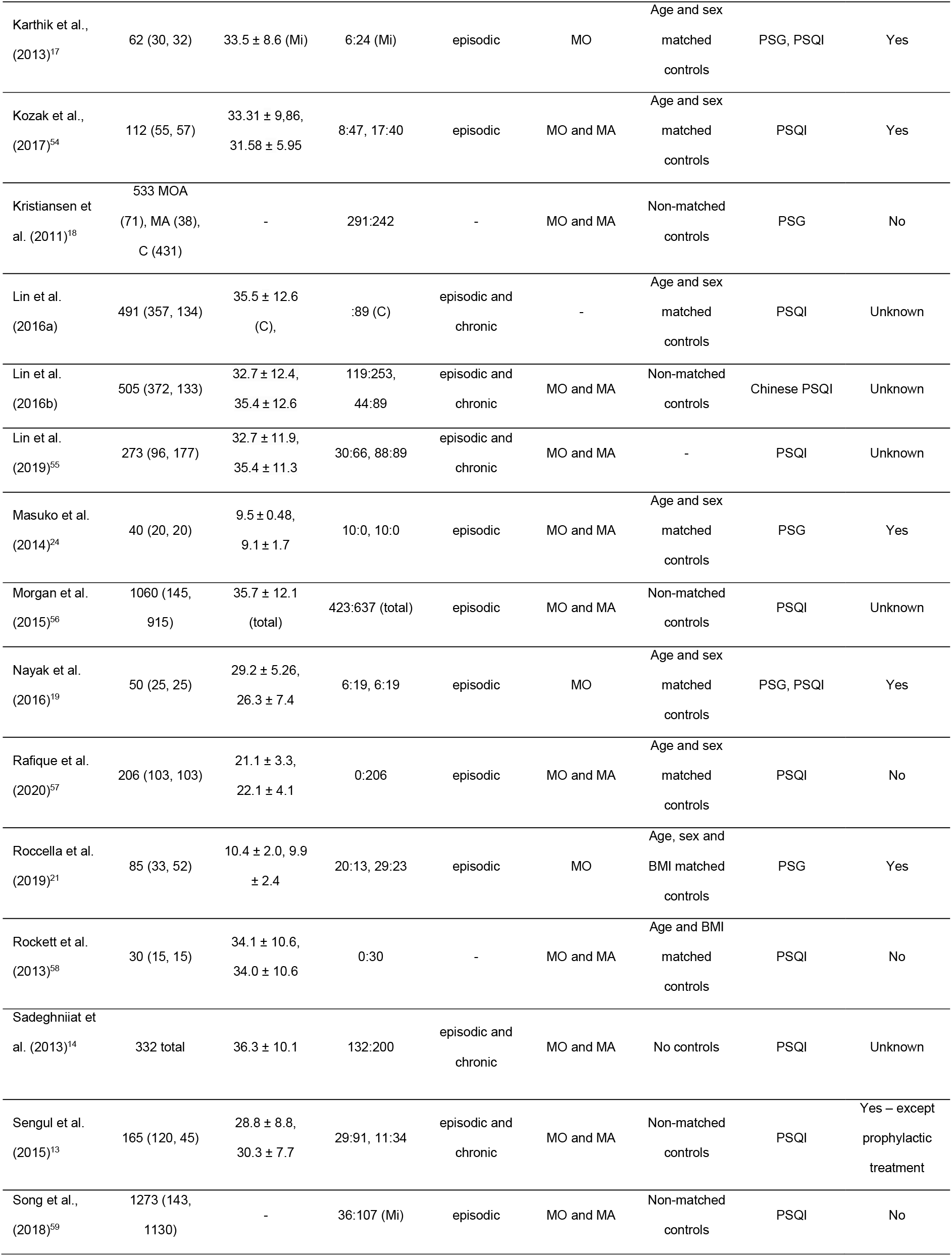
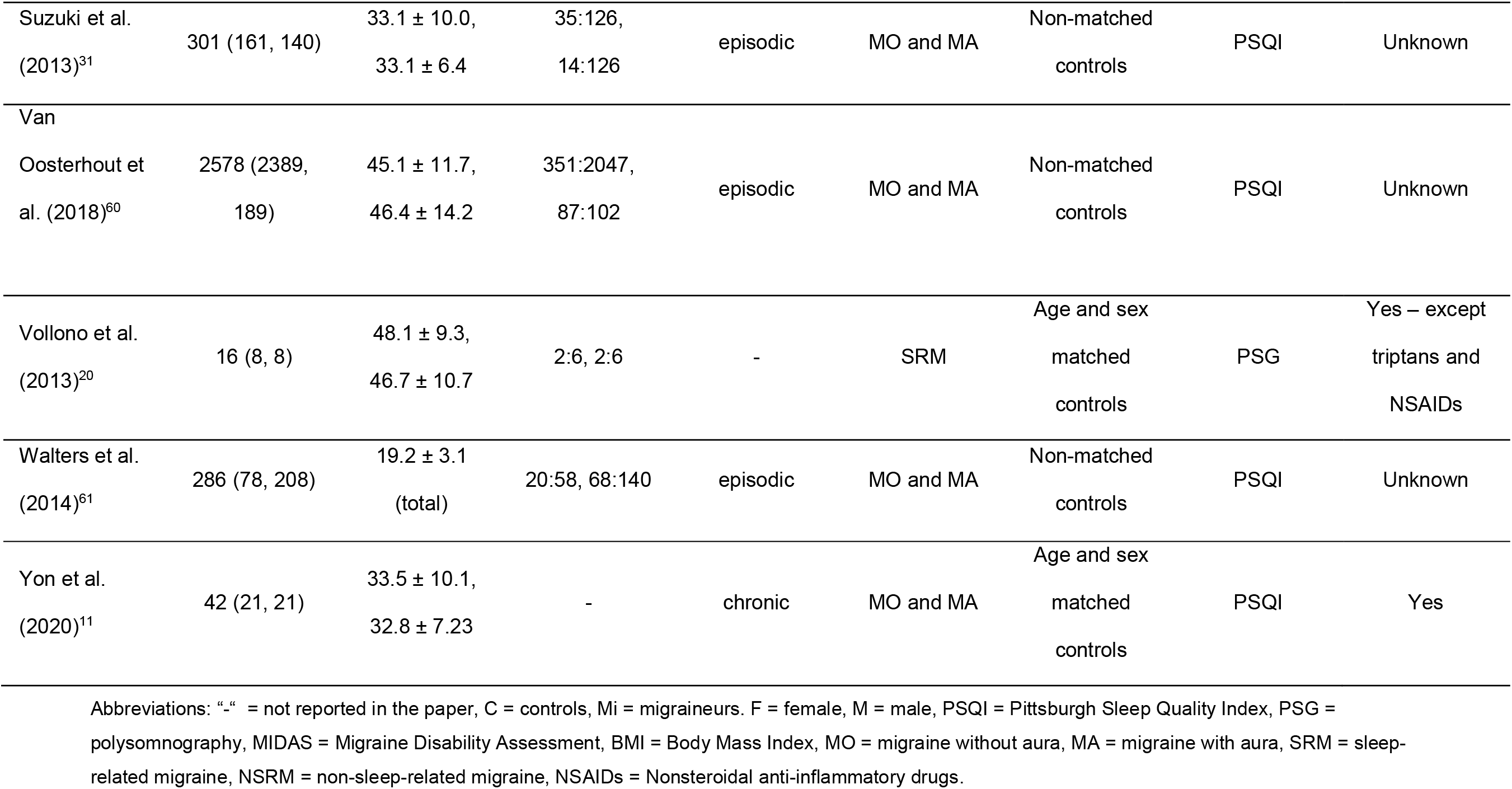
Methodological characteristics of the studies include in the meta-analysis

### 3.2. Risk of publication bias results

There was no indication of publication bias for any of the analyses apart from the PSQI and MIDAS correlation analysis as indicated by Egger’s test for funnel plot asymmetry (see Table e-3). The trim-and-fill method estimated that there was one study missing on the right-hand side, and this would increase the overall effect size to significance but not change the direction of effect (*z* = 0.44, *p* = 0.024). Although Egger’s test was not significant for any of the other analyses, visual inspection of the funnel plots was completed. For the PSG analysis, visual inspection, and the trim-and-fill method (see Figure e-1), revealed that for percentage wake there was likely to be one study missing on the right. However, adjustment of the effect size did not change the direction of overall effect or reduce the effect size, thus publication bias is unlikely to influence the result.

### 3.3. Meta-analysis results

#### 3.3.1. PSQI score

Figure 2. displays the forest plot of individual study-level as well as the overall effect size for the PSQI scores analysis in adult migraineurs and controls. Overall, there was a medium effect size for the difference in PSQI scores between migraineurs and healthy controls (*g* = 0.75, *p* < .001). For the chronic sub-group there was a large significant effect size (*g* = 1.03, *p* < .001) and for the episodic sub-group there was a medium effect size (*g* = 0.63, *p* < .001). The direction of these effect sizes indicates that migraineurs scored significantly higher on the PSQI than healthy controls, suggesting poorer sleep quality.

**Figure 2:**
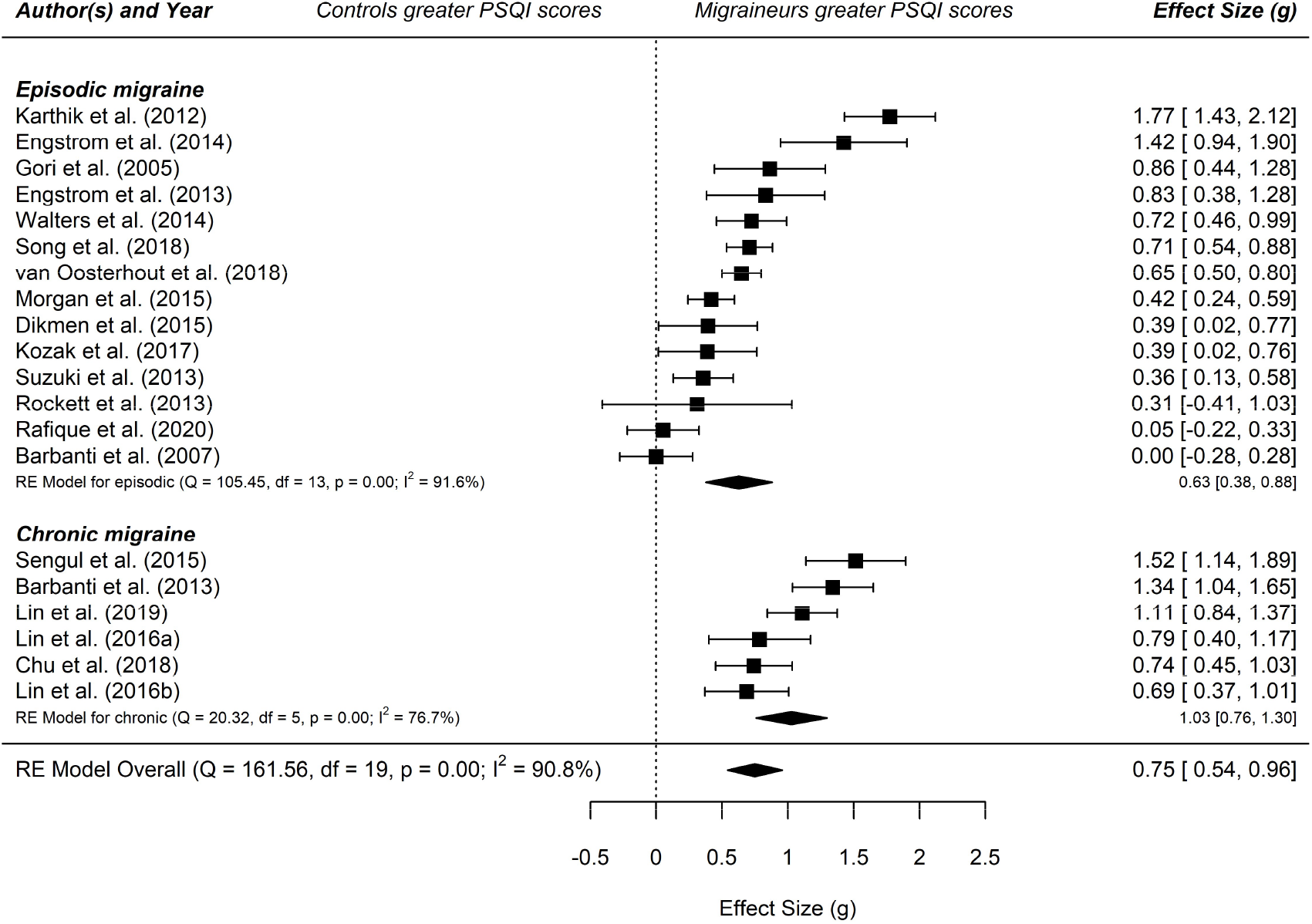
Forest plot of the meta-analysis of PSQI scores in migraineurs and controls. Abbreviations: PSQI, Pittsburgh Sleep Quality Index; RE, random effects. The standardized mean difference (Hedges’ *g*) and confidence intervals for the difference in global PSQI scores is shown between migraineurs and healthy controls.

For the overall analysis and the episodic subgroup there was statistically significant heterogeneity as evidenced by Cochran’s *Q* statistic (161.6 and 105.5, respectively). The chronic sub-group analysis had significantly lower heterogeneity (20.32) as measured by Cochran’s *Q*. However, when considering the *I*^2^ statistic the studies within the total, episodic, and chronic analyses all displayed moderate-high heterogeneity (*I*^2^ = 90.8%, 91.6%, 76.7% respectively), suggesting that over 70% of the variability is attributable to between-study heterogeneity, over and above sampling error.

As there was moderate or greater heterogeneity, moderator analyses were conducted on the overall analysis (see Table 4). There was a negative moderating effect of whether the study excluded those with sleep disorders or not (*Q*_*M*_ = 7.40, *p* = .007, *β* = −0.81), suggesting that when sleep disorders were excluded the effect size is smaller. However, there was significant heterogeneity not explained by this moderator (*Q*_*E*_ = 114.61, *p* < .001), and as only two studies excluded those with sleep disorders this could have skewed the result. No other variables were significant moderators.

#### 3.3.2. Polysomnography – adult & pediatric

Table 2 displays the effect sizes and heterogeneity measures for the PSG-derived sleep parameters in adult migraineurs and healthy controls. There was a significant small effect size for percentage REM sleep (*g* = −0.22, *p* = 0.017). The direction of this effect indicates that the adult migraineurs had less REM sleep than controls. There were no statistically significant effect sizes for the other parameters. There was significantly moderate heterogeneity as evidenced by the *Q* and *I*^2^ statistic in two of the analyses (SE and wake). However, as the main measure of effect size on these parameters was not significant no moderator analyses were conducted.

**Table 2:**
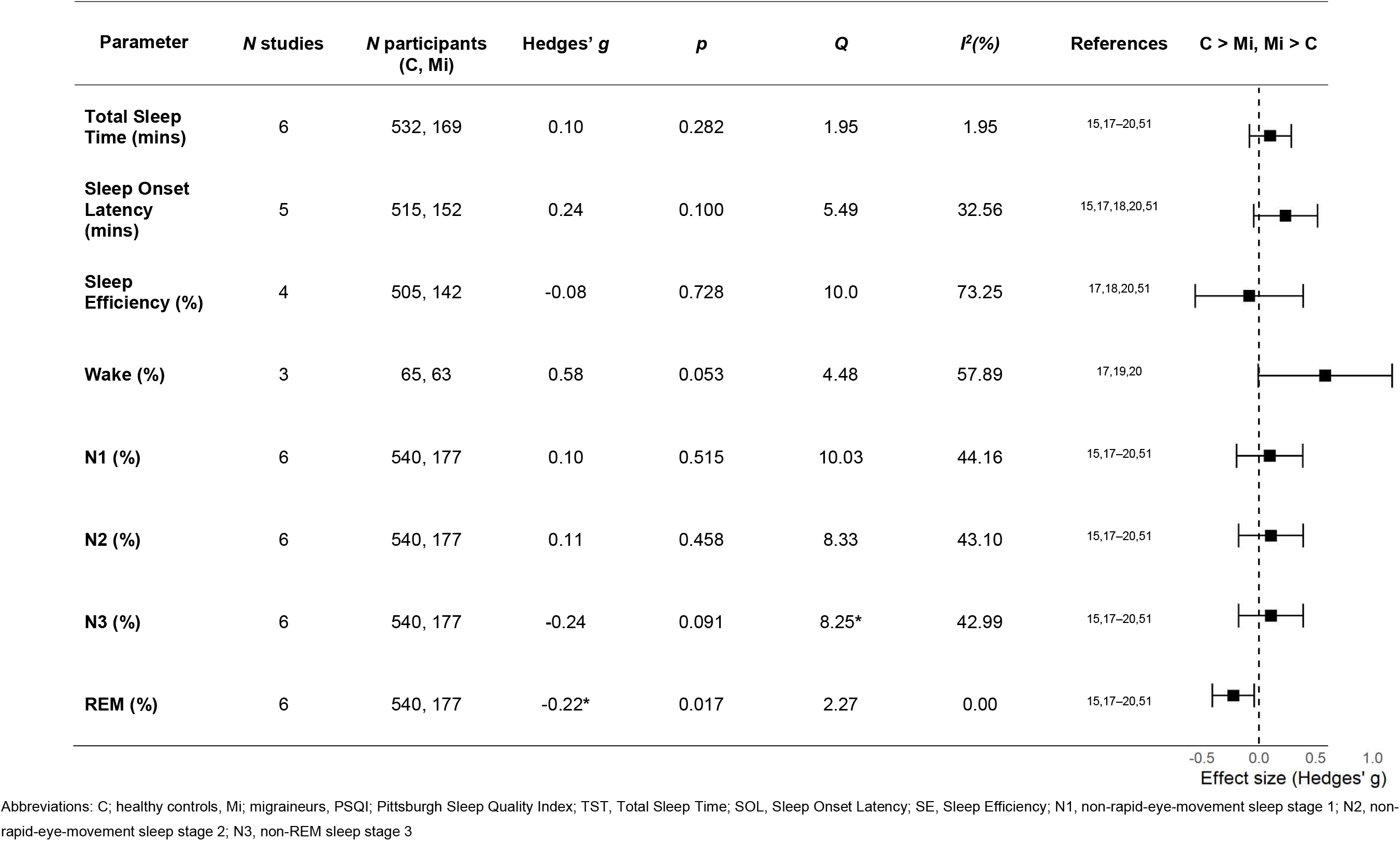
Results of the meta-analysis of PSG-derived sleep variables in adult migraineurs and control participants

| Parameter | N studies | N participants<br>(C, Mi) | Hedges' *g* | *p* | *Q* | *I*<sup>2</sup>(%) | References | C > Mi, Mi > C |
| --- | --- | --- | --- | --- | --- | --- | --- | --- |
| **Total Sleep Time (mins)** | 6 | 532, 169 | 0.10 | 0.282 | 1.95 | 1.95 | 15,17–20,51 |  |
| **Sleep Onset Latency (mins)** | 5 | 515, 152 | 0.24 | 0.100 | 5.49 | 32.56 | 15,17,18,20,51 |  |
| **Sleep Efficiency (%)** | 4 | 505, 142 | -0.08 | 0.728 | 10.0 | 73.25 | 17,18,20,51 |  |
| **Wake (%)** | 3 | 65, 63 | 0.58 | 0.053 | 4.48 | 57.89 | 17,19,20 |  |
| **N1 (%)** | 6 | 540, 177 | 0.10 | 0.515 | 10.03 | 44.16 | 15,17–20,51 |  |
| **N2 (%)** | 6 | 540, 177 | 0.11 | 0.458 | 8.33 | 43.10 | 15,17–20,51 |  |
| **N3 (%)** | 6 | 540, 177 | -0.24 | 0.091 | 8.25* | 42.99 | 15,17–20,51 |  |
| **REM (%)** | 6 | 540, 177 | -0.22* | 0.017 | 2.27 | 0.00 | 15,17–20,51 |  |

-0.5 0.0 0.5 1.0 Effect size (Hedges' *g*)
Abbreviations: C; healthy controls, Mi; migraineurs, PSQI; Pittsburgh Sleep Quality Index; TST, Total Sleep Time; SOL, Sleep Onset Latency; SE, Sleep Efficiency; N1, non-rapid-eye-movement sleep stage 1; N2, non-rapid-eye-movement sleep stage 2; N3, non-REM sleep stage 3

Table 3 displays the effect sizes for the PSG-derived parameters in pediatric migraineurs. There were small significant effect sizes for wake (*g* = 0.43, *p* = 0.015) and SOL (*g* = −0.37, *p* < .001). There was a medium significant effect size for REM sleep (*g* = −0.71, *p* = 0.025), and a large significant effect size for TST (*g* = −1.37, *p* = 0.039). The direction of these effects indicates that pediatric migraineurs had more wake, less REM sleep, less TST and longer SOL than healthy controls. There was statistically significant heterogeneity in six of the analyses (TST, SE, N1, N2, N3, REM).

**Table 3:** Results of the meta-analysis of PSG-derived sleep variables in pediatric migraineurs and control participant

| Parameter | *N* studies | *N* participants (C, Mi) | Hedges' *g* | *p* | *Q* | *I*<sup>2</sup>(%) | References | C > Mi, Mi > C |
| --- | --- | --- | --- | --- | --- | --- | --- | --- |
| **Total Sleep Time (mins)** | 4 | 146, 133 | -1.37* | 0.039 | 43.71* | 94.81 | 21–23,25 |  |
| **Sleep Onset Latency (mins)** | 4 | 303, 272 | -0.37* | <0.001 | 1.32 | 0.00 | 21,23–25 |  |
| **Sleep Efficiency (%)** | 5 | 323, 312 | -0.42 | 0.242 | 48.36* | 92.93 | 21–25 |  |
| **Wake (%)** | 3 | 123, 107 | 0.44* | 0.015 | 3.32 | 369.64 | 21–23 |  |
| **N1 (%)** | 5 | 323, 312 | -0.24 | 0.260 | 23.58* | 80.44 | 21–25 |  |
| **N2 (%)** | 5 | 323, 312 | 0.19 | 0.448 | 41.68* | 85.35 | 21–25 |  |
| **N3 (%)** | 5 | 323, 312 | 0.32 | 0.524 | 113.03* | 96.2 | 21–25 |  |
| **REM (%)** | 5 | 323, 312 | -0.71* | 0.025 | 32.0* | 90.42 | 21–25 |  |

-2 -1 0 1 Effect size (Hedges' *g*)
Abbreviations: C; healthy controls, Mi; migraineurs; PSQI; Pittsburgh Sleep Quality Index; TST, Total Sleep Time; SOL, Sleep Onset Latency; SE, Sleep Efficiency; N1, non-rapid-eye-movement sleep stage 1; N2, non-rapid-eye-movement sleep stage 2; N3, non-REM sleep stage 3

**Table 4:** Results of the moderator analysis of global PSQI scores in adult migraineurs

| Analysis | $Q_M$ | $P$ | $\beta$ | $Q_E$ | $p$ |
| --- | --- | --- | --- | --- | --- |
| **Sleep disorders excluded** | 7.40* | 0.007 | -0.81 | 114.61* | <.001 |
| **Matched controls** | 0.02 | 0.878 | 0.03 | 161.41* | <.001 |
| **Medications excluded** | 0.32 | 0.572 | 0.13 | 160.34* | <.001 |

Abbreviations: PSQI = Pittsburgh Sleep Quality Index

Two of the analyses of PSG variables in pediatric migraineurs which found significant effect sizes and displayed at least moderate heterogeneity were: TST and REM, thus a moderator analysis was conducted on these. None of the studies excluded patients with sleep disorders, so this was not included as a moderating variable. Table 5 displays the results. For TST, both whether the study excluded patients who were on sleep-affecting medications, and whether the study included matched controls or not, were significant moderating variables (*Q*_*M*_ = 19.6, *p* < .001, *Q*_*M*_ = 19.6, *p* < .001) respectively. The test for residual heterogeneity (*Q*_*E*_) was not significant for these two analyses, indicating that they are largely influencing the effect size in these studies. The direction of this moderator analysis indicated that when a study did exclude those on medication, or did include matched controls, the effect size was smaller. However, whether the study included an adaptation night or not was not a significantly moderating variable (*Q*_*M*_ = 0.20, *p* = 0.655, Q_*E*_ = 38.46, *p* < .001). For REM sleep, there were no significant moderators of effect size and *Q*_*E*_ was significant for all analyses, indicating significant residual heterogeneity that cannot be explained by these moderators, and thus other variables are influencing between-study heterogeneity.

**Table 5:** Results of the moderator analysis of PSG-derived sleep variables in pediatric migraineurs

| Analysis | $Q_M$ | $p$ | $\beta$ | $Q_E$ | $p$ |
| --- | --- | --- | --- | --- | --- |
| **Adaptation night** |  |  |  |  |  |
| *TST* | 0.20 | 0.655 | -0.69 | 38.46* | < .001 |
| *REM* | 0.01 | 0.931 | 0.10 | 28.37* | < .001 |
| **Matched controls** |  |  |  |  |  |
| *TST* | 19.60* | <.001 | -2.51 | 4.75 | 0.093 |
| *REM* | 0.80 | 0.372 | -0.70 | 17.73* | < .001 |
| **Medications excluded** |  |  |  |  |  |
| *TST* | 19.60* | <.001 | -2.51 | 4.75 | 0.093 |
| *REM* | 0.80 | 0.372 | -0.70 | 17.73* | < .001 |

Abbreviations: TST = total sleep time, REM = rapid-eye-movement sleep, PSG = polysomnography

#### 3.3.3. Relationship between sleep quality and migraine disability

Figure 3. shows the weighted effect size for the correlation between MIDAS and PSQI scores in migraineurs. There was a small non-significant effect size for the correlation between MIDAS scores and PSQI scores in migraineurs (*z* = 0.32, *p* = 0.060), thus suggesting from this analysis that there is no relationship between sleep quality and migraine disability. There was significantly low heterogeneity between studies as indicated by Cochran’s *Q* statistic (*Q* = 28.74, *p* < .001), however *I*^2^% indicated high heterogeneity (*I*^2^% = 91.1%), and as this statistic is more appropriate for small samples, high heterogeneity between these studies can be assumed. As the main analysis was not significant no moderator analyses were conducted.

**Figure 3:**
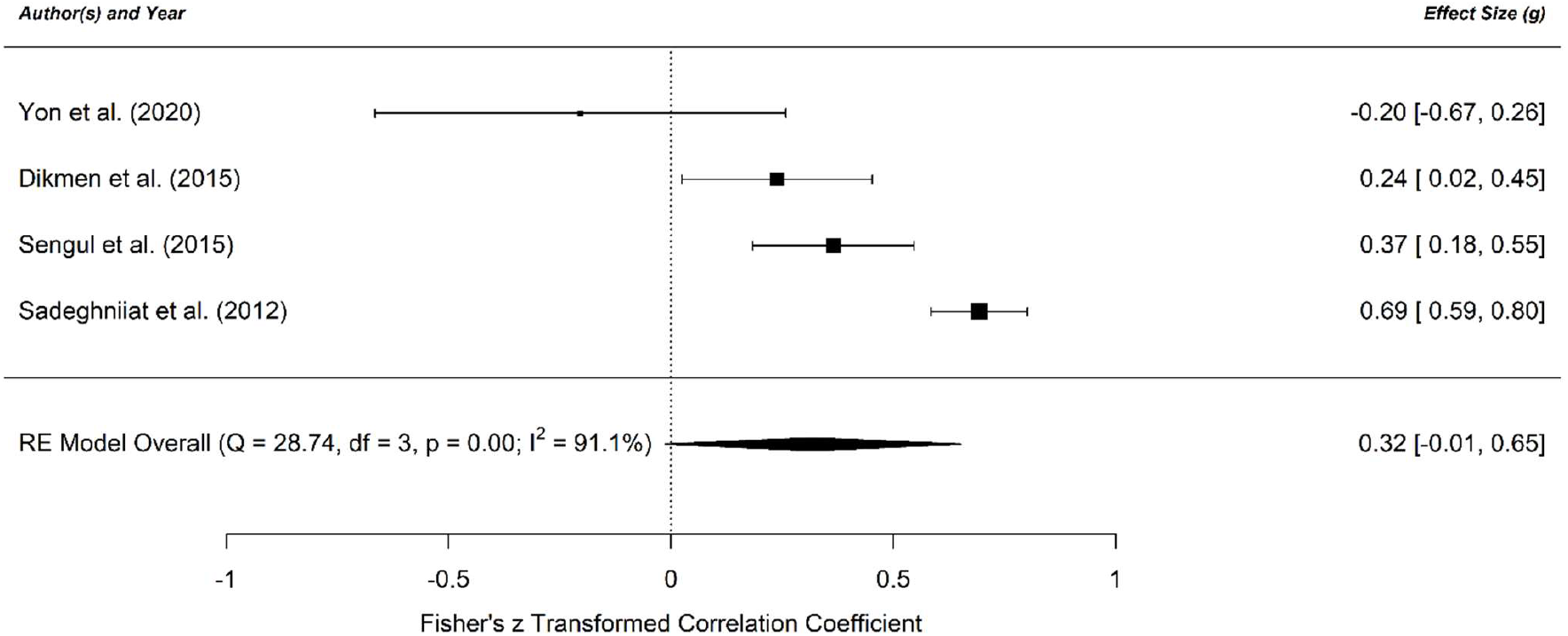
Forest plot of the meta-analysis of MIDAS and PSQI correlations. Abbreviations: RE, random effects. The Fisher’s z transformed correlation coefficient is shown between MIDAS scores and PSQI scores in patients with migraine across four studies.

## 4. Discussion

This meta-analysis aimed to consolidate previous findings and establish whether there are differences in subjective sleep quality and objective sleep physiology between migraineurs and healthy individuals. The findings demonstrate that migraineurs display significantly higher scores on the PSQI indicating worse subjective sleep quality than healthy controls, an effect which is larger in those with chronic rather than episodic migraine. There are differences in objective sleep physiology, although this is mainly evident in the pediatric population. For example, adult and pediatric migraineurs have significantly less REM sleep as a percentage of TST than healthy individuals, and this effect size is larger in the pediatric population. Pediatric migraineurs also exhibit significantly less TST, shorter SOL, and more wake than healthy controls. There were no differences between migraineurs and healthy controls in any other sleep parameters either in adult or pediatric populations. Finally, there was no significant overall correlation between PSQI and MIDAS scores in migraineurs.

### 4.1. Subjective sleep quality

These findings extend the current literature by aggregating the results of multiple studies, thereby increasing power. They demonstrate that sleep quality is worse in migraineurs than healthy individuals. Although the PSQI may be criticized as a subjective measure of sleep, it has been shown to have high test-retest reliability and high validity^26^. Moreover, this finding is not surprising; poor sleep is reported as an exacerbating factor for migraines in 50% of cases^27^. The effect size was larger in chronic migraineurs. This is in line with previous research where poor sleep has been shown to be an important factor in progression to chronic migraine^28^.

### 4.2. Polysomnography

PSG on the other hand provides a more objective measure of sleep. This analysis found that REM sleep is reduced in both adult and pediatric migraineurs, relative to healthy individuals, aligned with previous associations between migraine and REM sleep. For example, a co-association between narcolepsy type 1 (NT1) and migraine has been demonstrated^29^ and migraine is reported as a risk factor for progression to NT1 in children^30^. Moreover, one study reported an association between dream enactment behavior during REM sleep and migraine^31^. Furthermore, cutaneous allodynia, a prominent symptom experienced during migraine attacks, has been shown to worsen in response to REM sleep deprivation in preclinical pain models^32^. This indicates a potential dysfunction in mechanisms underlying REM-NREM or REM-wake transitions in migraineurs. A plausible neural correlate for this is the hypothalamic orexinergic system which plays a critical role in stabilizing sleep/wake transitions and REM sleep^34^, and has been linked to migraine, suggesting multiple points of intersection between sleep and trigeminal pain. Although, this is speculative, and the relationship with REM sleep is likely to be complex and bi-directional. For example, some preventative treatments for migraine can suppress REM sleep whilst others lead to increased arousals during REM.

There were no differences between adult migraineurs and controls in any other sleep parameters. This is at odds with a previous meta-analysis which found that TST, WASO and SOL were worse in those with chronic pain (including migraineurs) than healthy controls^10^. Although, this might be reconciled in that the number of studies they included was larger and contained heterogeneous chronic pain conditions. It is likely that with a greater number of studies, we might see significant differences in wake, TST, and SOL in adults.

Conversely, pediatric migraineurs displayed lower TST, lower REM sleep percentage, and more wake in line with previous findings in pediatric patients with chronic pain^35^. The migraineurs also displayed shorter SOL than controls. This is suggestive of pediatric migraineurs operating at higher sleep pressure due to being chronically sleep deprived (perhaps due to headache, behavior, or both) hence their shorter SOL times. Alternatively, migraineurs may be biologically sleepier than their healthy counterparts. Their shorter SOL is at odds with an actigraphy study in pediatric migraineurs which found a longer SOL in patients compared to healthy controls^36^. However, actigraphy is widely considered to be less accurate than PSG, particularly for SOL^37^.

The lack of any significant difference in NREM sleep, including N3, in both adults and children with migraine compared to controls is noteworthy, although this represents macroscopic sleep EEG scoring rather than quantitative EEG analysis. Indeed, deficits in NREM sleep can be compensated for by an increase in sleep intensity rather than duration as is the case typically for REM sleep^38^, which would not be apparent with macroscopic scoring.

### 4.3. Migraine disability

The lack of a significant correlation between PSQI and MIDAS scores is surprising given that previous studies report significant correlations between migraine-related disability and sleep disturbances^39^, and altering sleep (e.g. by switching back to day shifts from night shifts) can have a profound effect on migraine-related disability scores^40^. This may reflect the small number of studies and the high heterogeneity between them. Three of the studies in this analysis found significant positive correlations^14^, and one found a large negative correlation^11^. Studies with a small *n* can lead to inflated effect sizes^41^. Indeed, the study which found a negative relationship had an *n* of 21^11^, compared to 332 for one of the larger studies, thus potentially skewing the results towards insignificance. There was also evidence of publication bias within this analysis. Although, caution should be taken when interpreting the results as despite meta-analysis being theoretically sound on a small number of studies, Egger’s test can produce false positives.

### 4.4. Limitations

A critical consideration is that many migraineurs are given prophylactic treatments which can affect the sleep cycle. When this factor was included as a moderating variable in the pediatric analysis for TST and REM sleep, studies which excluded those on medications had a smaller effect size for TST than those which did not, suggesting that the patient’s medication may be contributing to differences in TST. Indeed, beta-blockers – a common migraine preventative, are known to reduce TST. Nonetheless, significant residual heterogeneity remained between studies which could not be explained by this moderator, suggesting other factors are at play.

Importantly, medication exclusion did not impact REM sleep. Although, it is unclear whether medication was driving the REM sleep effect seen in adults as no moderator analysis was conducted on this. Furthermore, for many studies it was impossible to deduce whether patients on medication were excluded. Future studies should ensure accurate reporting of exclusion criteria.

These results do not provide evidence for a direct functional relationship between migraine and sleep regulation. Are migraineurs experiencing poor sleep due to the pain caused by attacks, or are they experiencing attacks due to poor sleep? The studies in this analysis did not report whether the migraineurs typically experience their migraine attacks during sleep itself, despite two thirds of migraineurs reporting experiencing them during sleep^27^. Only a handful of studies reported whether the PSG was conducted in the ictal or inter-ictal period, despite this being shown to have a considerable effect on objective measures of sleep^42^. If this were to affect the results however, we might have expected to see a difference in other sleep parameters in both the adults and pediatric migraineurs. As we only see the decrease specific to REM sleep and no other sleep stages, this implies a specific dysfunction in the mechanisms that underly REM sleep.

Alternatively, the reduced REM sleep could reflect the finding that many migraine attacks occur in the early hours of the morning -the portion of the night where REM sleep dominates, hence curtailing REM opportunity. However, two of the PSG studies in the current analysis had a population of migraineurs with predominantly sleep-related attacks^16,20^ and neither study reported differences in REM sleep between SRM, NSRM or controls. This implies that the decrease in REM sleep we see here is not necessarily due to arousals from REM sleep during an attack.

Moreover, six of the included studies did not include an adaptation night to the sleep laboratory. The ‘first night effect’ has been shown to particularly affect measures of REM sleep^43^. However, the moderator analyses in pediatric migraineurs found that whether the study included an adaptation night or not was not a significant moderator of the overall effect size for REM sleep or TST.

The disparity between the large effect size seen with the subjective measure of sleep and the lack of significant difference between migraineurs and controls with all but one of the objective sleep physiology parameters in the adult population is not unexpected. Global PSQI scores have been shown not to significantly correlate with sleep variables as measured via PSG^44^. As we did not analyze PSQI scores in the pediatric population, it is not clear if pediatric migraineurs also experience altered subjective sleep quality. This being said, the PSQI has been shown to have limited utility in pediatrics^45^.

In addition, many of the studies were retrospective studies of patients who have previously been referred to the sleep clinic with non-specific sleep complaints, necessitating PSG assessment, suggesting the migraineurs referred may already report underlying sleep complaints. Few of the studies mentioned prior sleep history in relation to PSG, and for subjective sleep quality those which did exclude sleep disorders had a smaller effect size than those which did not, suggesting that poor sleep quality may be attributable to undiagnosed concurrent sleep disorders.

However, this is unlikely to be the case as the heterogeneity between studies was not fully explained by this variable. This residual heterogeneity may reflect differences in pre-recording sleep history, differing recording environments, techniques and equipment, as well as differences in inter-scorer scoring concordance^46^.

### 4.5. Clinical implications

These findings highlight that consideration of patient sleep should play an integrated role in the assessment and treatment of migraine. Clinicians should consider prioritizing behavioral sleep interventions (especially in children with migraine) as well as considering sleep when prescribing medication. Recent studies have demonstrated the utility of sleep behavioral interventions in reverting chronic to episodic migraine^47^, which may suggest a potential role of instigating similar interventions earlier in the natural history of the disorder to mitigate the risk of conversion from episodic to chronic migraine. The relevance of REM sleep and its modulation for migraine and migraine treatment is again emphasized by this meta-analysis. The relationship is likely to be complex and related to sleep (and indeed REM) homeostasis, rather than related to any absolute proportions of REM sleep, a notion supported by the REM suppressing (rather than enhancing) effects of the best-known migraine preventive, amitriptyline.

### 4.6. Conclusions

Migraineurs report poorer subjective sleep quality than healthy individuals, an effect larger in those with chronic migraines. Adult migraineurs exhibit significantly less REM sleep compared to healthy controls. Pediatric migraineurs also show significantly reduced sleep time, faster sleep onset and more wake than controls. The interplay between migraine and sleep is likely to be highly complex and remains poorly understood. However, the findings of this meta-analysis highlight the importance of assessing and treating sleep in migraineurs as an integrated part of headache treatment. While offering significant insight into how sleep is relevant to migraine, this study also highlights the limitations of drawing conclusions from small case-controlled PSG studies of patients with migraine given the significant number of confounds and heterogeneity involved. Future studies may do well to analyze the interplay between migraine and subjective sleep factors at large-scale population-based levels, as well as also using a more population-based approach to sleep physiological studies, performed in a standardized way, to minimize variability where possible.

## Data Availability

The source data are available from the individual studies included. The extracted raw data are available at Open Science Framework.

https://osf.io/3t4u5/

## 5. Appendix - Authors

| Name | Location | Role | Contribution |
| --- | --- | --- | --- |
| Emily C. Stanyer, MPsych | King's College London, United Kingdom | Author | Designed and conceptualized the study; collected, performed statistical analyses, interpreted the data; and drafted and revised the manuscript for intellectual content |
| Hannah Creeney, MSc | King's College London, United Kingdom | Author | Major role in the acquisition of data |
| Alexander D. Nesbitt, BM BCh, PhD | Guy's and St Thomas' NHS Foundation Trust, London, United Kingdom | Author | Revised the manuscript for intellectual content |
| Philip R. Holland, PhD | King's College London, United Kingdom | Author | Revised the manuscript for intellectual content |
| Jan Hoffmann, MD, PhD | King's College London, United Kingdom | Author | Designed and conceptualized the study; interpreted the data; revised the manuscript for intellectual content |

