## Supplementary Figure 1 for "Subjective sleep quality and objective sleep physiology in migraineurs: a meta-analysis"

**Figure e-1:** Trim-and-fill funnel plots for each of the meta-analyses in migraineurs and controls

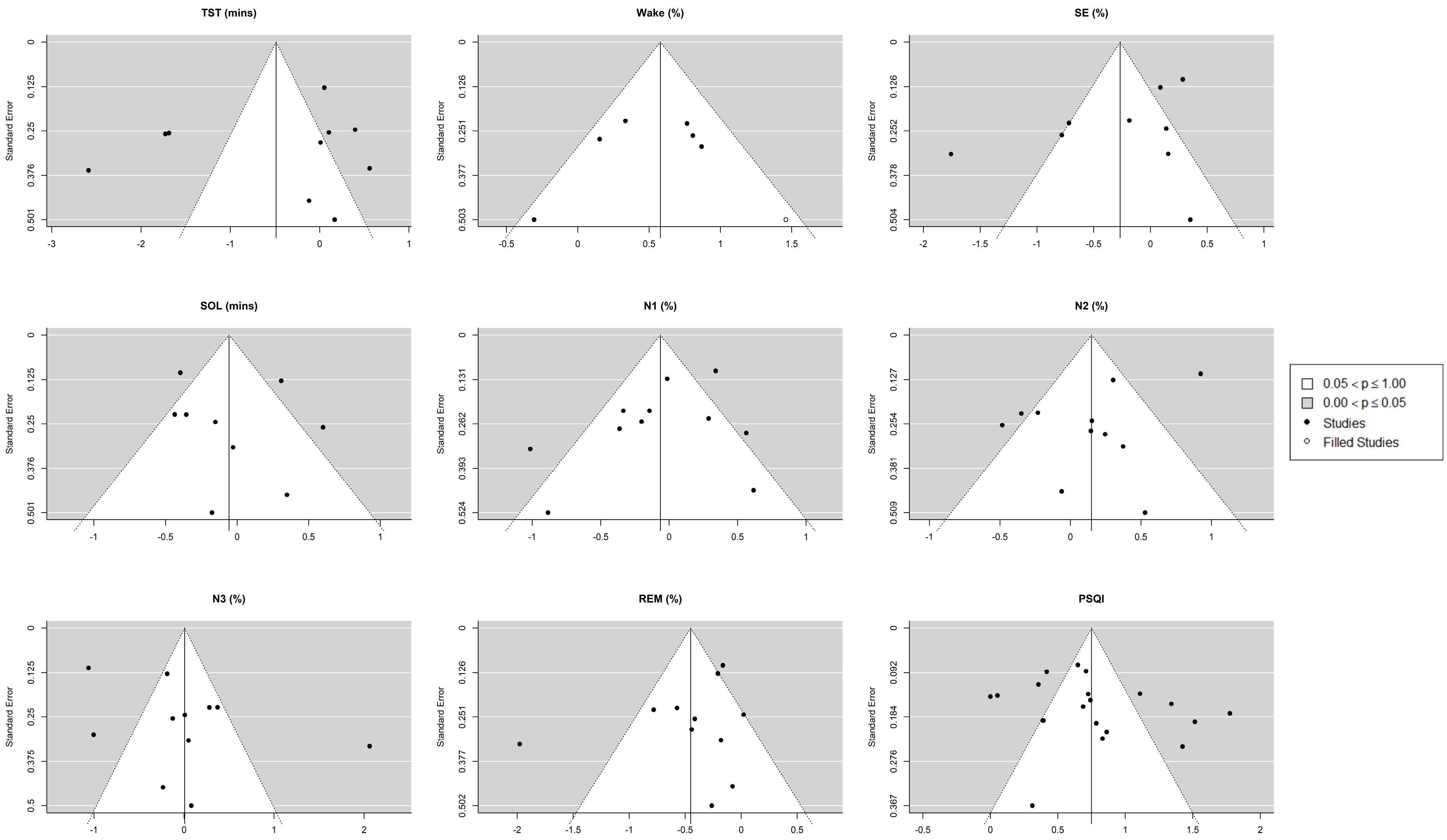


Abbreviations: PSQI; Pittsburgh Sleep Quality Index; TST, Total Sleep Time; SOL, Sleep Onset Latency; SE, Sleep Efficiency; N1, non-rapid-eye-movement sleep stage 1; N2, non-rapid-eye-movement sleep stage 2; N3, non-REM sleep stage 3; mins = minutes.
