## Supplementary Table 2 for "Subjective sleep quality and objective sleep physiology in migraineurs: a meta-analysis"

**Table e-2:** Search strategy for OVID

| Search line number | Keyword/s | Search string | Number of results |
|---|---|---|---|
| **Databases selected:** Embase (1974 – 2020), Ovid MEDLINE (R) (1946 – 2020), Global Health (1973 – 2020), APA PsycInfo (1806 – 2020), APA PsycArticles Full Text (- 2020) | | | |
| 1 | migraine* | mp=ti, ab, hw, tn, | 128610 |
| 2 | MIDAS | mp=ti, ab, hw, tn, | 4457 |
| 3 | Migraine Disability | mp=ti, ab, hw, tn, | 2154 |
| 4 | MS-Q | mp=ti, ab, hw, tn, | 286 |
| 5 | HIT-6 | mp=ti, ab, hw, tn, | 1283 |
| 6 | Headache Impact Test | mp=ti, ab, hw, tn, | 1441 |
| 7 | sleep quality | mp=ti, ab, hw, tn, | 63290 |
| 8 | PSQI | mp=ti, ab, hw, tn, | 12845 |
| 9 | Pittsburgh Sleep Quality | mp=ti, ab, hw, tn, | 22948 |
| 10 | EEG | mp=ti, ab, hw, tn, | 244685 |
| 11 | electroencephalograph* | mp=ti, ab, hw, tn, | 333289 |
| 12 | PSG | mp=ti, ab, hw, tn, | 19344 |
| 13 | polysomnograph* | mp=ti, ab, hw, tn, | 82692 |
| 14 | 1 or 2 or 3 or 4 or 5 or 6 | | 131646 |
| 15 | 7 or 8 or 9 | | 63520 |
| 16 | 10 or 11 or 12 or 13 | | 500081 |
| 17 | 15 or 16 | | 552637 |
| 18 | 14 and 17 | | 4934 |
| 19 | Limit 18 to English language | | 4155 |
| 20 | Remove duplicates from 19 | | 4089 |
