## Supplementary Table 3 for "Subjective sleep quality and objective sleep physiology in migraineurs: a meta-analysis"

**Table e-3:** Results of Egger's regression test for publication bias

| Analysis | *z* | *p*_value |
|---|---|---|
| **PSQI** | -0.771 | 0.441 |
| **TST (mins)** | 0.059 | 0.953 |
| **Wake (%)** | -1.275 | 0.202 |
| **SOL (mins)** | 0.501 | 0.617 |
| **SE (%)** | -0.451 | 0.652 |
| **N1 (%)** | -1.080 | 0.280 |
| **N2 (%)** | 1.262 | 0.207 |
| **N3 (%)** | 0.881 | 0.378 |
| **REM (%)** | -0.504 | 0.614 |
| **MIDAS & PSQI correlations** | -4.33* | <.001 |

Abbreviations: REM, rapid-eye-movement sleep; TST, total sleep time; PSQI, Pittsburgh Sleep Quality Index; SOL, Sleep Onset Latency; SE, Sleep Efficiency; N1, non-rapid-eye-movement sleep stage 1; N2, non-rapid-eye-movement sleep stage 2; N3, non-REM sleep stage 3. * = statistically significant at $p < 0.05$.
